# Seizure Semiology and Response to Treatment in a Pediatric Cohort with *SCN2A* Variants: A Parent Report

**DOI:** 10.1101/2023.02.23.23286378

**Authors:** John B. O’Connor, Emily B. Kirschenblatt, Linda Laux, Anne T. Berg, Sunita N. Misra, John J. Millichap

## Abstract

We examined seizure semiology and response to medications in 28 children (9 female, 19 male) with likely pathogenic and pathogenic *SCN2A*-related epilepsy. Parents reported seizure onset, seizure semiology, genetic variants, therapies used for seizures, and response to treatment. 27 children experienced defined seizures and 1 reported no seizure history. The most common initial seizures were focal or hemi-convulsions (n=8). Tonic seizures were the most common reported seizure type while febrile and atonic or drop seizures were the least common. Most patients experienced multiple seizures daily or were entirely seizure-free, with no difference based on age at seizure onset. The proportion of effective trials of the 8 most commonly reported medications ranged from 4 of 26 trials (levetiracetam) to 5 of 10 trials (valproic acid). Phenytoin was the most commonly reported effective treatment (N=4). Topiramate was reported to be the most effective treatment in combination with another treatment (N=6). We found a wide phenotypic spectrum of *SCN2A*-related disorders and a possible correlation between genotype and seizure onset, semiology, and treatment response. Gain-of-function mutations in early-onset *SCN2A* epilepsies responded well to sodium channel blockers. Further exploration of *SCN2A* pathogenic variants are needed to identify mediation mechanisms of action in *SCN2A*-related epilepsy.

## INTRODUCTION

*SCN2A* encodes one of the major α subunits, Na_v_1.2, of voltage-gated sodium channels in neurons^1^. *De novo* pathogenic or likely pathogenic variants in *SCN2A* cause various symptoms, and variants have been reported in patients with epileptic encephalopathies, autism spectrum disorder (ASD), and intellectual disability with epilepsy^2-5^.

Symptoms associated with *SCN2A*-related epilepsy, including recurrent unprovoked seizures and cognitive impairment, can range from mild to severe and life-threatening^6^. Early treatment can help prevent developmental delay and allow for effective control of seizures^7,8^; however, current treatment approaches rely on empirical techniques, and little is known about the causal electrophysiological mechanisms of these complex disorders. As is the case with other complex neurodevelopmental brain disorders, a better understanding of treatment response in *SCN2A*-related epilepsies can inform the selection of more effective treatments earlier in the disease course, which can mitigate devastating consequences such as impaired brain development, the inability to walk or communicate, and early mortality^7,8^.

A previous report described that earlier-onset *SCN2A*-related epilepsies are associated with gain-of-function variants, while later-onset epilepsies are seen in children with loss-of-function variants^10^. As a result, sodium channel blockers are expected to be more effective in children with earlier-onset seizures, and less effective or even harmful in children with later-onset seizures^10^. A more comprehensive understanding of genotype-phenotype correlations regarding seizure type and response to medication is needed to improve neurodevelopmental outcomes in children with *SCN2A*-related disorders. The objective of this study was to describe seizure semiology and the response to various medications in children with *SCN2A* variants based on parent report. Responses for the entire study cohort were analyzed, and comparisons were made by variant and age at seizure onset.

## RESULTS

### Cohort demographics

A total of 36 participants with presumed *SCN2A* variants were recruited to the study; of those, 33 provided clinical data by parental report, including 28 who also provided genetic reports or medical records confirming the variant. Analyses were performed on data for the 28 participants for whom both clinical and genetic data were made available. Participants were 0.90 to 15.24 years of age (mean=5.48 years, standard deviation=3.58 years) at the time of parent report. Nine (32.1%) participants were female, and 19 (67.9%) were male, 26 (92.9%) were Caucasian, and 2 (7.1%) were Asian. Three (10.7%) identified as Hispanic, 24 (85.7%) identified as non-Hispanic, and 1 (3.6%) did not indicate their ethnicity.

### Genotype

The variants of the 28 participants are outlined in Table 1. There were 28 distinct variants, with one recurrent variant (p.R853Q) in two separate individuals and two variants (p.K1260E and p.K1260Q) in one individual. The two variants identified in the same participant were found at the same locus, indicating that the participant exhibited compound heterozygosity, which is representative of atypical autosomal recessive inheritance. The 28 distinct variants included 2 splice site variants, 25 missense variants(including the compound heterozygous variants), and 1 deletion insertion (Supplemental Figure 1). Of the 26 variants not predicted to impact splicing (25 missense and 1 deletion-insertion), 1 (4%) was in the first domain, 2 (8%) were in the domain 1-domain 2 linker region, 6 (23%) were in the second domain, 3(12%) were in the domain 2-domain 3 linker region, 5 (19%) were in the third domain (2 from the compound heterozygous participant), 1 (4%) was in the domain 3-domain 4 linker region, 4 (15%) were in the fourth domain, and 4 (15%) were in the C-terminal region of the protein subunit. Of these variants, 1 (4%) was extracellular, 11 (42%) were in transmembrane locations including the pore domain, and 14 (54%) were cytoplasmic (2 from the compound heterozygous participant).

**Table 1:**
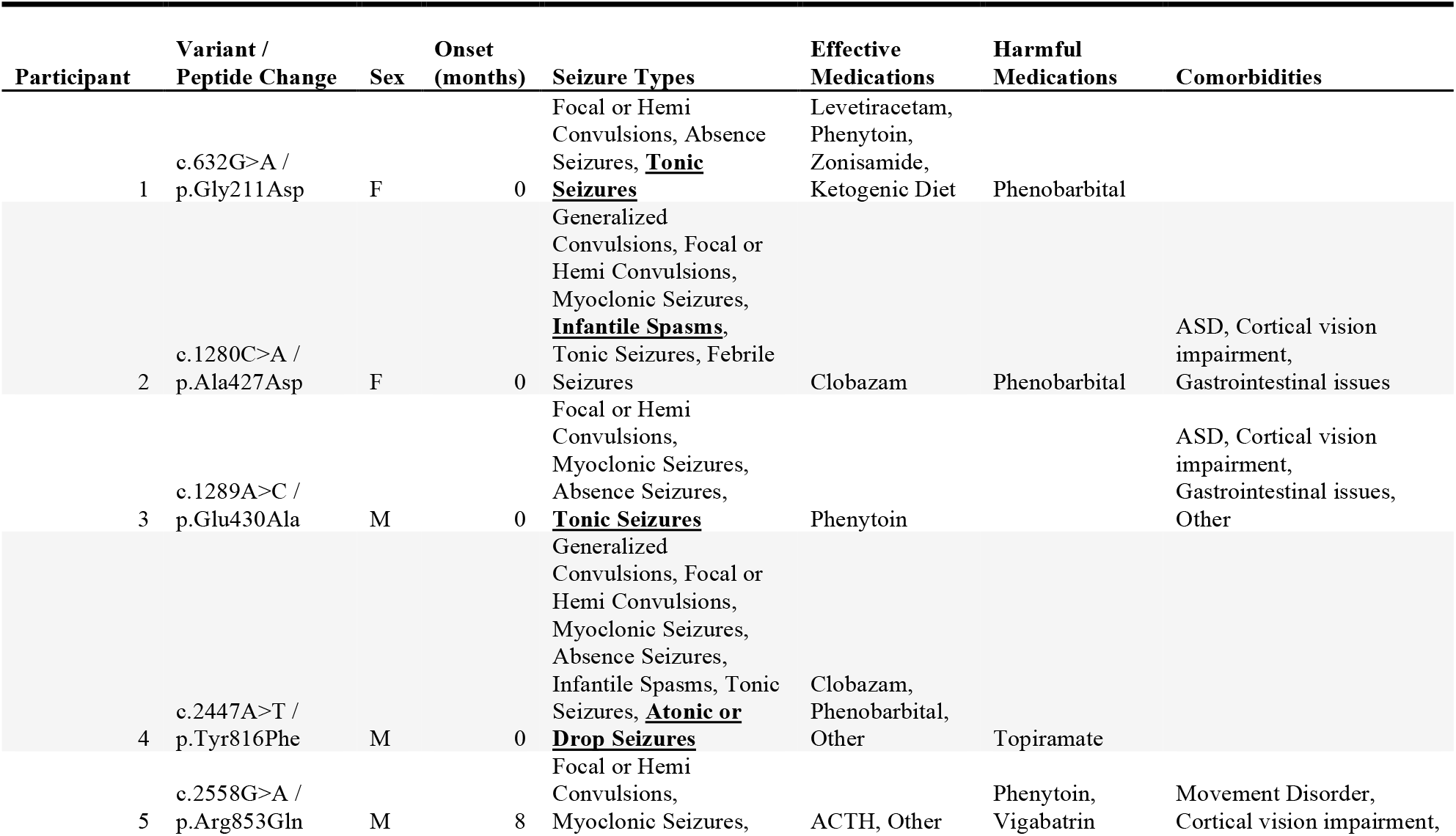
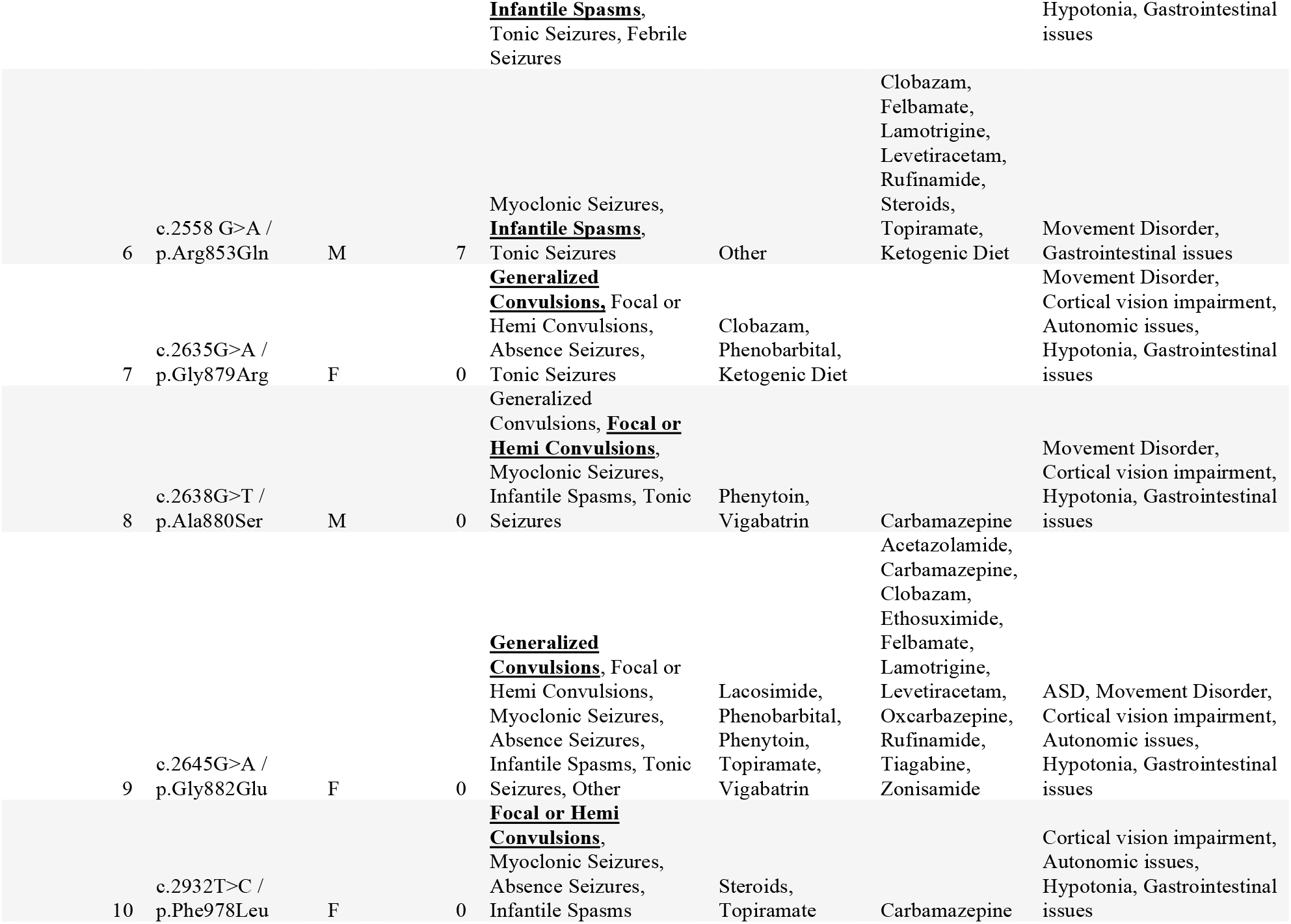
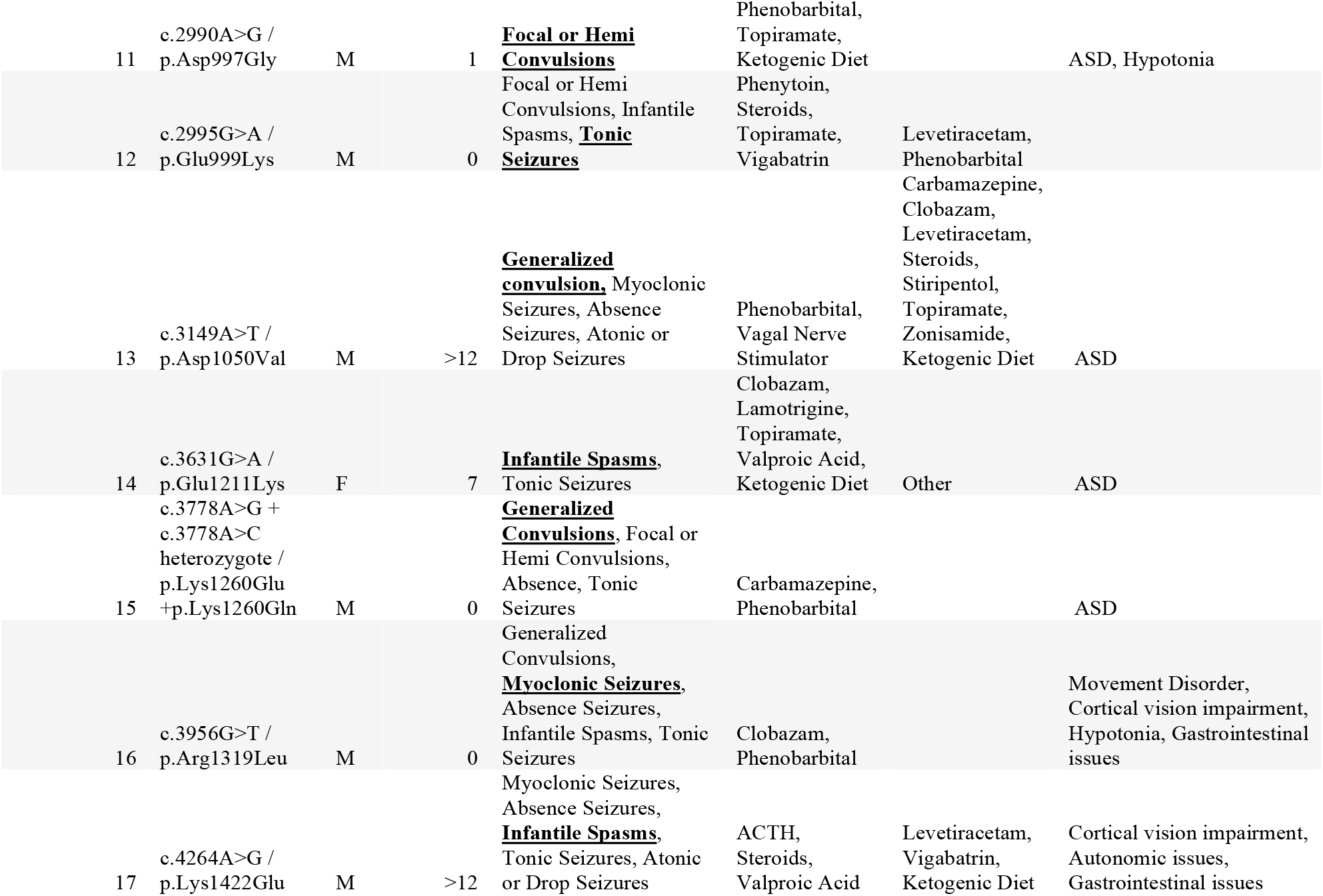
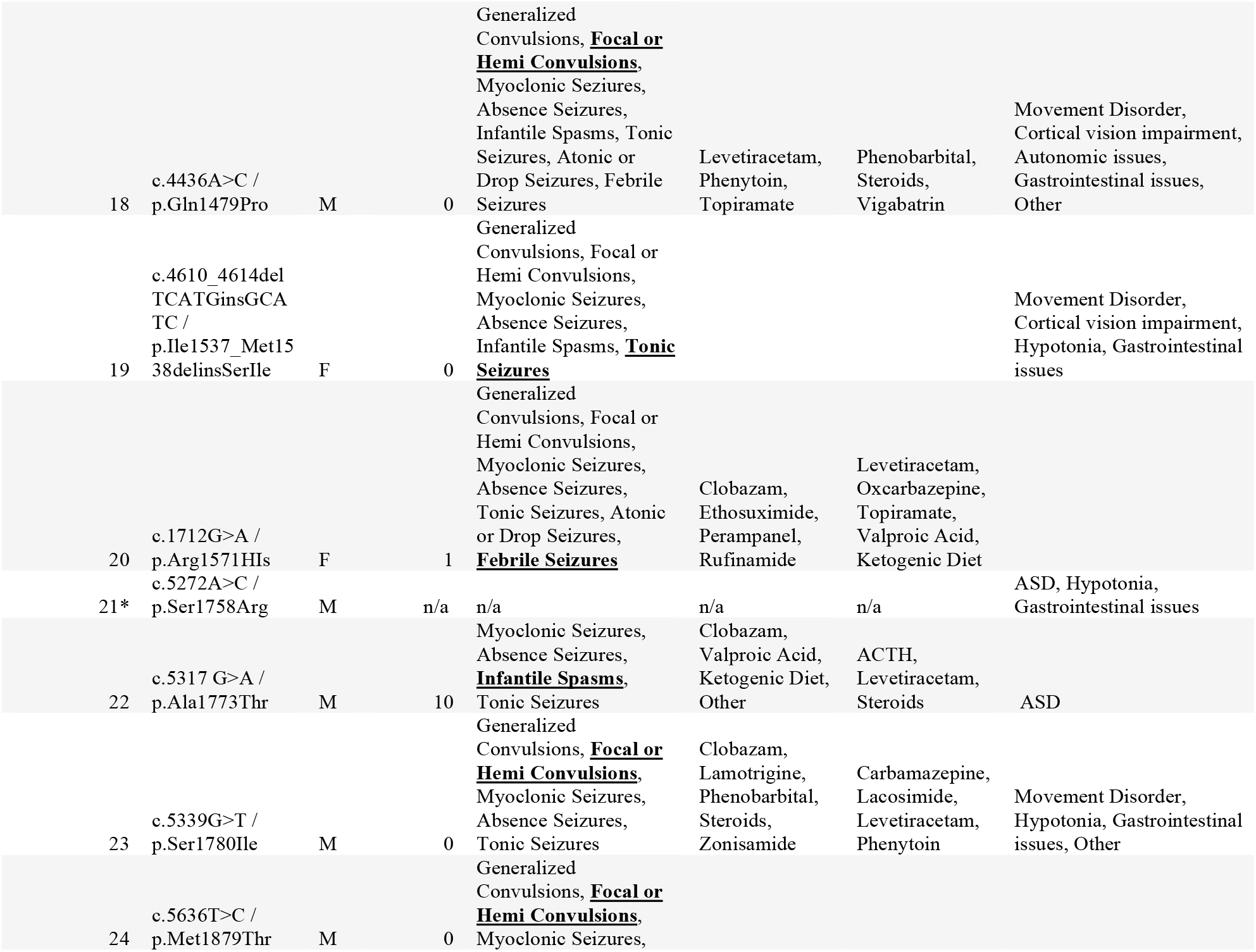
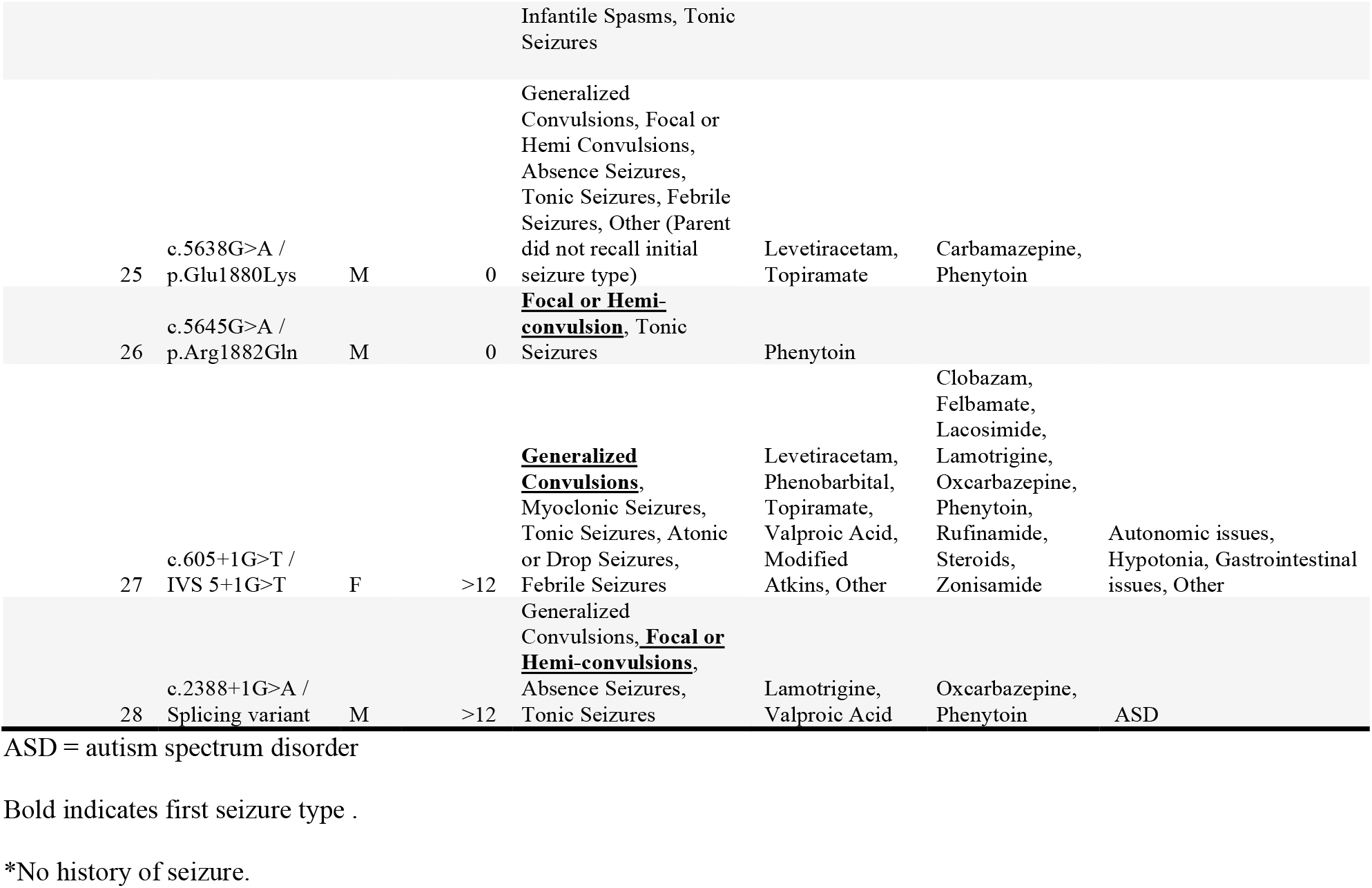
Summary of Genotype, Seizure Onset, Seizure Semiology, and Response to Medications in the Study Cohort

Further analysis of the 26 missense or deletion insertion variants revealed that 3 (12%) were in the voltage sensor region (S4), 7 (27%) were in the pore domain (S5, S5-S6 linker, and S6), and 1 (4%) was within the fast inactivation gate (3–4 domain linker). Furthermore, 2 variants, p.A427D and p.E430A, were reported in the 1–2 domain linker, which is a region sensitive to phosphorylation-dependent modulation of channel activity^11^. Six variants (23%) were also located in toxin binding sites.

### Seizure onset

The parent of 1 participant (4%) did not report a defined seizure onset due to no history of seizures, 17 (61%) reported neonatal onset, 2 (7%) reported onset at 1 month of age, 2 (7%) at 7 months, 1 (4%) at 8 months, 1 (4%) at 10 months, and 4 (14%) after the first year of life. Of those with a defined seizure onset, 23 (85%) reported an onset within the first year of life.

Of the 27 participants with defined seizure onset, the initial seizure type was reported to be an atonic or drop seizure in 1 (4%) participant, febrile seizure in 1 (4%) participant, focal or hemi-convulsions in 8 (30%) participants, generalized convulsions in 5 (19%) participants, infantile spasms in 6 (22%) participants, myoclonic seizure in 1 (4%) participant, and tonic seizures in 4 (15%) participants (Figure 1). One parent did not recall their child’s initial seizure onset. Of the 17 participants with neonatal onset, the more common initial reported seizure types were focal or hemi-convulsions (n=6, 35%), tonic seizures (n=4, 24%), and generalized convulsions (n=3, 18%). Of the 4 participants with onset after 1 year of age, the most common reported seizure type was generalized convulsions (n=2, 50%).

**Figure 1:**
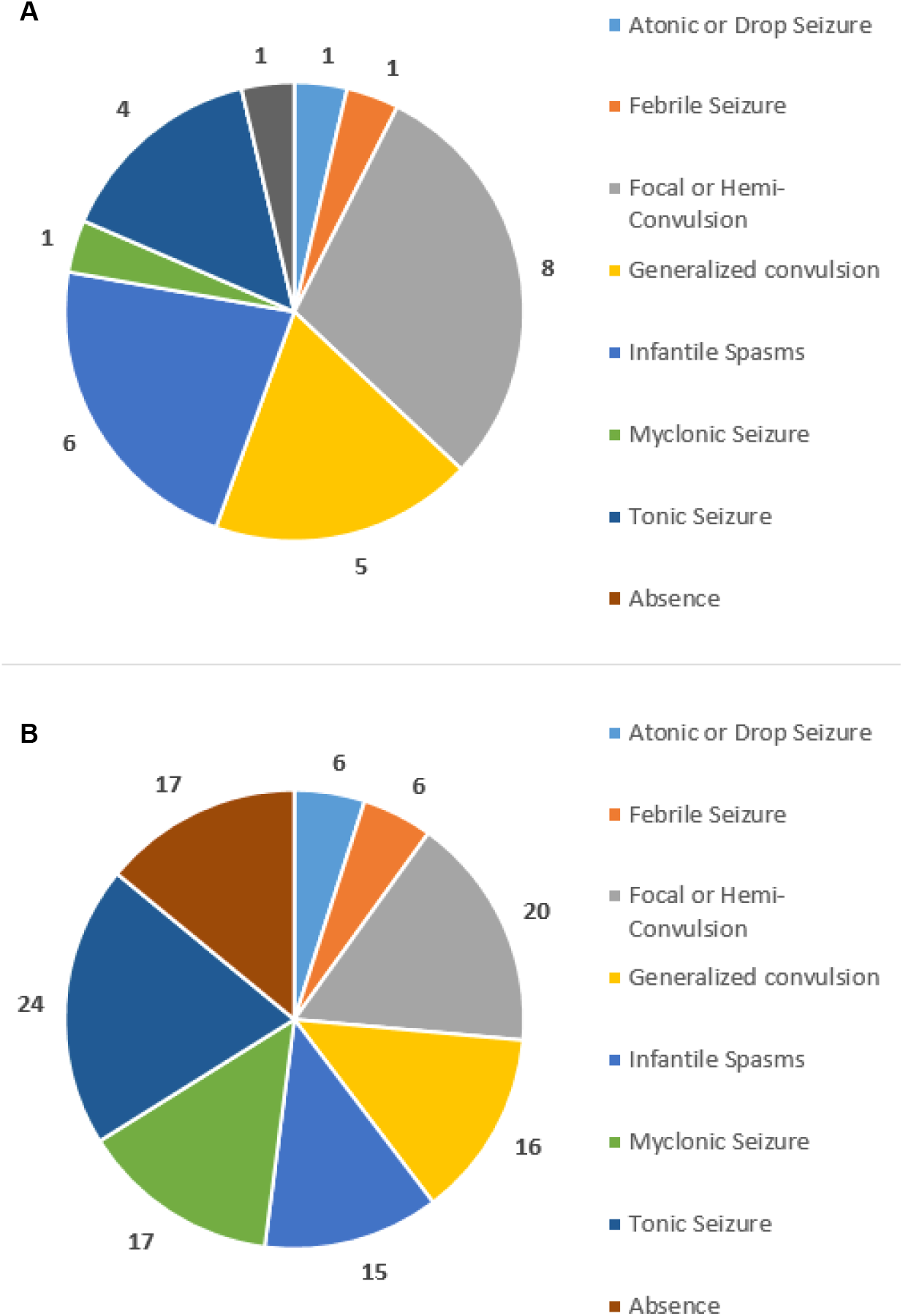
**(A)** Distribution of seizure type at initial onset in 27 patients. (**B)** Distribution of seizure type across time in 27 patients with reported initial onset seizures.

When asked whether or not the first seizure was triggered by any event, 18 (67%) reported that the initial seizure was unprovoked, 2 (7%) reported that the initial seizure was due to an illness with fever, 1 (4%) reported the initial seizure was due to vaccination, 4 (15%) reported some other cause, and 2 (7%) did not provide a response. According to parental report, 22 (81%) had an initial seizure that was under 5 minutes, 2 (7%) were between 5 and 30 minutes,1 (4%) was over 30 minutes, and 2 (7%) had an unknown initial duration.

### Seizure semiology

When the parents who reported seizure onset (N=27) were asked about a history of other seizures in their child, responses ranged from 0 to 7 other seizure types, with a median of 4 seizures reported (IQR=4 to 6). The prevalence of the different reported seizure types is summarized in Figure 2. Tonic seizures were the most common, reported in 24 (89%) cases. Of the known seizure types, febrile and atonic seizures were the least common, reported in 6 (22%) cases. For the participant with the p.R1319L variant, myoclonic seizures were not reported in the overall history; however, that seizure type was reported at seizure onset and was included in the participant’s overall seizure history. Comparisons between the seizure distribution at initial onset and seizure distribution across time in the 27 participants are shown in Figure 2.

**Figure 2:**
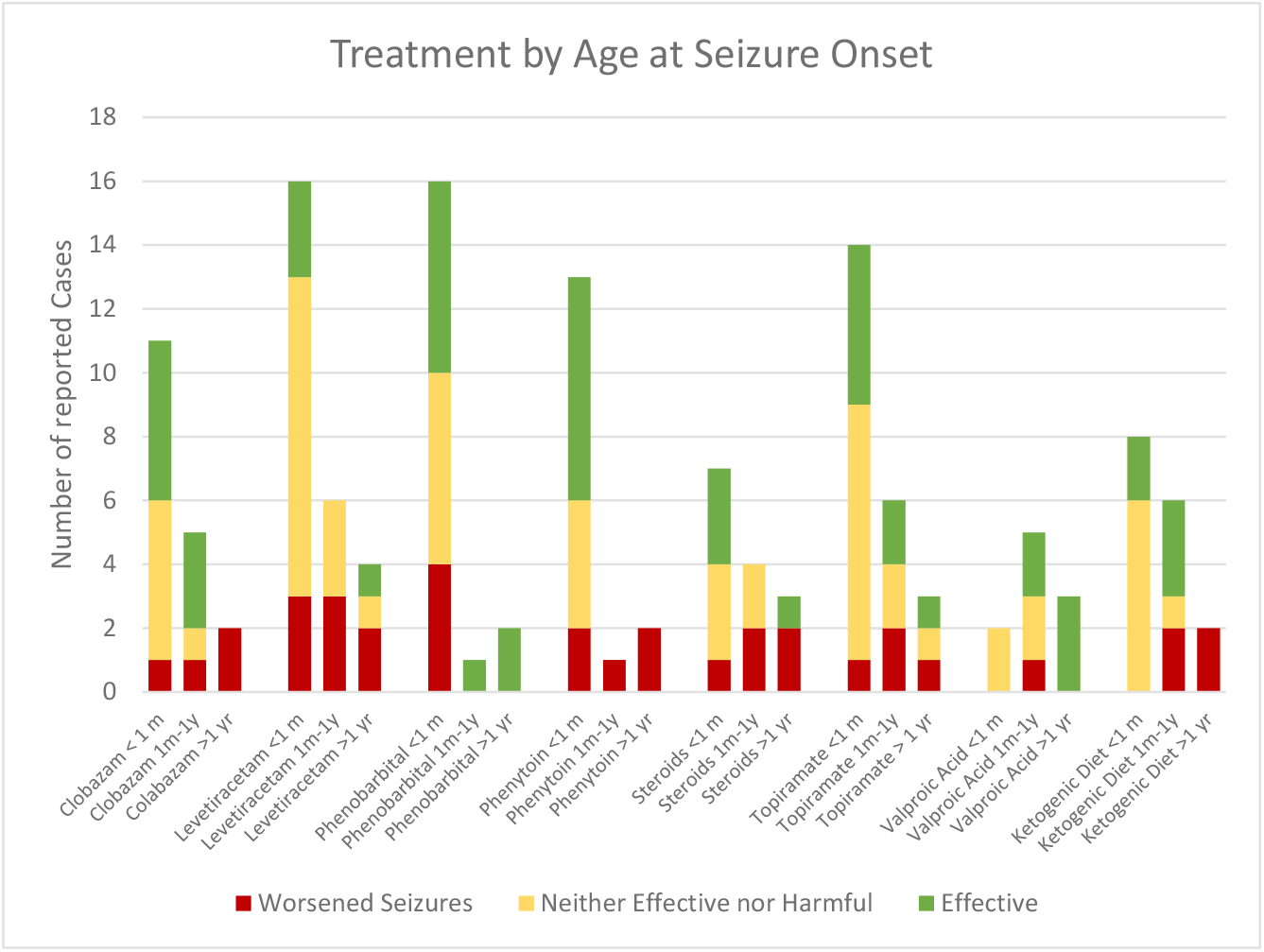
Counts of parent-reported responses separated by age at seizure onset.

When asked about seizure frequency within the last two months, parents reported that the majority of participants were either seizure-free (7, 26%) or experienced multiple seizures a day (9, 33%). Two (7.4%) reported rare frequency, 1 (3.7%) reported monthly seizures, 5 (18.5%) reported weekly seizures, and 3 (11.1%) reported daily seizures. The same seizure frequency proportions were seen when comparing participants with earlier- and later-onset seizures (Figure 2).

Parents were also asked how the frequency of seizures compared to the child’s typical seizure control to provide context. Of the 7 participants who were seizure-free in the previous 2 months, 1 (14%) parent described it as “typical” and 6 (86%) described it as “good.” Of the 9 participants who had multiple daily seizures, 2 (22%) parents described it as “good,” 2 (22%) described it as “typical,” and 5 (55%) described it as “poor.” When asked about the longest time that their child was reported to be seizure-free, 2 (7%) did not provide a response, 3 (11%) indicated days, 4 (15%) indicated weeks, 11 (41%) indicated months, and 4 (26%) indicated years.

### Drug response

As expected, the 1 participant without any defined seizures had not trialed any of the antiepileptic treatments included in the survey. Of the 27 parents of children with a reported seizure history, all indicated trialing multiple treatments, ranging from 4 to 25 different treatments with a median number of 8 (IQR=6 to 11). The 8 most commonly trialed treatments were levetiracetam (n=26, 96.3%), topiramate (n=24, 88.8%), clobazam (n=20, 74.1%), phenobarbital (n=19, 70.3%), the ketogenic diet (n=17, 63.0%), phenytoin (n=16, 59.3%), steroids (n=14, 51.9%), and valproic acid (n=12, 44.4%). None of the parents reported a trial of eslicarbazepine or ezogabine. When analyzing seizure response, 6 responses were removed from analysis due to ambiguity. The responses are summarized in Supplemental Figure 2. Beneficial responses for the commonly trialed treatments ranged from 15% (levetiracetam) to 50% (valproic acid). Comparisons between patients with neonatal onset versus onset after 1 year of age suggested potential differences in drug response by age at onset (Figure 2).

Parents were also asked about a single effective therapy that helped their child the most. Of the 27 parents of children with reported seizures, 19 (70%) indicated that there was one medication that helped their child the most, 6 (22%) replied that there was no single most effective therapy, and 1 (7%) did not provide a response. The results of the single most effective treatments are summarized in Supplemental Table 1.

Parents were also asked about which medications worked in combination (Supplemental Table 2). Of the 27 participants who had a seizure history, 8 (30%) did not select any effective combinations. One response, indicating only carbamazepine was an effective combination, was omitted due to ambiguity. The combinations included in the analysis varied, with responses ranging from only 2 to a combination of 5 treatments (clobazam, lamotrigine, topiramate, valproic acid, ketogenic diet). The median number of medications included in a combination was 2 (IQR=2 to 3). Clobazam and topiramate were the two most common treatments found to be effective in combination, reported in 3 (11%) participants.

### Autism spectrum disorder and other comorbidities

When asked whether their child was diagnosed with autism spectrum disorder (ASD), 10 (36%) parents indicated that their child was diagnosed, 17 (61%) that their child was not diagnosed, and 1 (4%) did not provide a response. ASD was found to be less prevalent in participants with earlier-onset epilepsies, reported in only 4 of the 17 neonatal onset participants but 5 of the 9 participants with onsets at 1 month and older. These findings, however, were not statistically significant (p=0.1025).

## DISCUSSION

In this study, parent-reported responses demonstrated the phenotypic heterogeneity of *SCN2A-*related disorders. The children in our cohort had diverse seizure semiology, with most parents indicating that their child had multiple seizure types. Tonic and focal or hemi-convulsions were reported to be the most prevalent, while atonic and febrile seizures were indicated to be the least frequent in this cohort. When assessing seizure frequency, we found most parents reported that either their child had multiple seizures daily or was completely seizure-free. Of those who indicated that their child was-seizure free, most described it as a good outcome; of those who indicated that their child was experiencing multiple daily seizures, most described it as a poor outcome.

As in previously published analyses of SCN2A variants, we found a possible connection between genotype, initial seizure onset, and overall seizure history. Participants with the recurrent p.R853Q variant had a similar onset age (7 and 8 months). Additionally, the first seizure type reported for both individuals with the recurrent p.R853Q variant was infantile spasms. These results, in addition to the initial seizure type in the participant with the p.E1211K variant and the onset ages of the participants with the p.E430A and IVS 5+1G>T variants, are consistent with the results reported by Wolf et al.10, indicating the possibility of a correlation between initial seizure type and underlying pathogenic variant. When comparing later seizures that occurred after onset, similar genotype-phenotype correlations were observed. The two participants with the p.R853Q variant both experienced myoclonic seizures and tonic seizures in addition to infantile spasms at initial onset. A history of spasms and/or tonic seizures were also reported by Ben-Shalom et al. in 4 of 5 participants with p.R853Q variants12. Additionally, myoclonic seizures, infantile spasms, and tonic seizures were also experienced by the participant in our cohort with variant p. R1319L, in the fourth voltage-sensing segment. This suggests a potential correlation between variants in the voltage-sensing segments and seizure type. Overall, the reported seizure semiologies support the possibility of a correlation between seizure type and underlying pathogenic *SCN2A* variant.

Across our cohort, parent responses about medications indicated that most participants were trialed on multiple anti-epileptic treatments. Of those, valproic acid, phenobarbital, and clobazam were most effective in children with pathogenic *SCN2A* variants. Valproic acid was least likely to exacerbate seizures. While our small sample size precludes us from making strong inferences, preliminary comparisons suggest that certain treatments—specifically clobazam, phenytoin, and the ketogenic diet—might be more effective for children with neonatal seizure onset who tend to have gain-of-function mutations, while phenobarbital and valproic acid might have greater efficacy in children with later onset and loss-of-function variants. The findings for phenytoin are consistent with those of Howell et al., who demonstrated that phenytoin can be at least partially effective for seizure control in early-onset *SCN2A* epilepsies^13^.

There also appears to be a correlation between response to medications and functional effects of SCN2A variants. The variant p.R1882Q has been previously reported to be gain-of-function, and the recurrent p.R853Q variant has been reported to be loss-of-function^14^. The participant with p.R1882Q responded well to phenytoin, a sodium channel blocker. In contrast, one of the participants with the recurrent loss-of-function variant reportedly experienced seizure exacerbation with phenytoin, while the other participant with a loss-of-function mutation was reported to experience worsening of seizures with lamotrigine, another sodium channel blocker. Similarly, the participant with the p.E1211K variant, which was reported to be gain-of-function^12^, responded well to lamotrigine. Further exploration into the functional effects of *SCN2A* pathogenic variants could identify the mechanisms by which some of these medications control seizures in *SCN2A*-related epilepsy. This functional understanding could inform treatment selection earlier on, potentially limiting the severity of negative outcomes, such as impaired brain development, the inability to walk or communicate, or early mortality.

## METHODS

Parents of children with *SCN2A* variants were recruited to take part in a study within Lurie Children’s Epilepsy Center, which was approved by the Institutional Review Board (IRB) of Ann & Robert H. Lurie Children’s Hospital of Chicago. Parents were provided with an IRB-approved study information sheet, and, due to the study’s low-risk designation, consent was obtained verbally from parents according to all IRB rules and regulations. After parents provided verbal consent to participate in the study, they provided a genetic report or medical records indicating their child’s *SCN2A* genetic variant. Information regarding seizure onset, seizure type, trialed medications, and response to those medications, was collected through parent report.

All data were collected through a secured REDCap survey and exported to Microsoft Excel. Any data with conflicting responses were excluded from analysis. Basic descriptive and bivariate statistical techniques were used to analyze the response data of the study population including frequency percentages for categorical variables. For statistical comparisons of categorical variables, Fisher’s exact tests were performed in SAS.

## Data Availability

All data produced in the present work are contained in the manuscript.

## ACKNOWLEDGEMENTS

The authors acknowledge the FamiliesSCN2A organization for their support and help in study promotion and subject recruitment. The authors also acknowledge the contribution of collaborators at Northwestern University, specifically Dr. Alfred George, in study conception and design.

## AUTHOR CONTRIBUTIONS

JMM and ATB conceived the study; JBO, EB, and JMM enrolled patients; JBO, SRM, JMM, and ATB consolidated and analyzed data, JBO, JMM, SRM, EB, and ATB wrote the manuscript, and all authors reviewed the manuscript.

## COMPETING INSTERSTS STATEMENT

Dr. Millichap reports royalties from Up-To-Date; consulting fees from Esai, Xenon, Biomarin, Greenwich, Praxis, Neurelis, Neurocrine, Biohaven.

## SUPPLEMENTARY INFORMATION

**Supplemental Figure 1:**
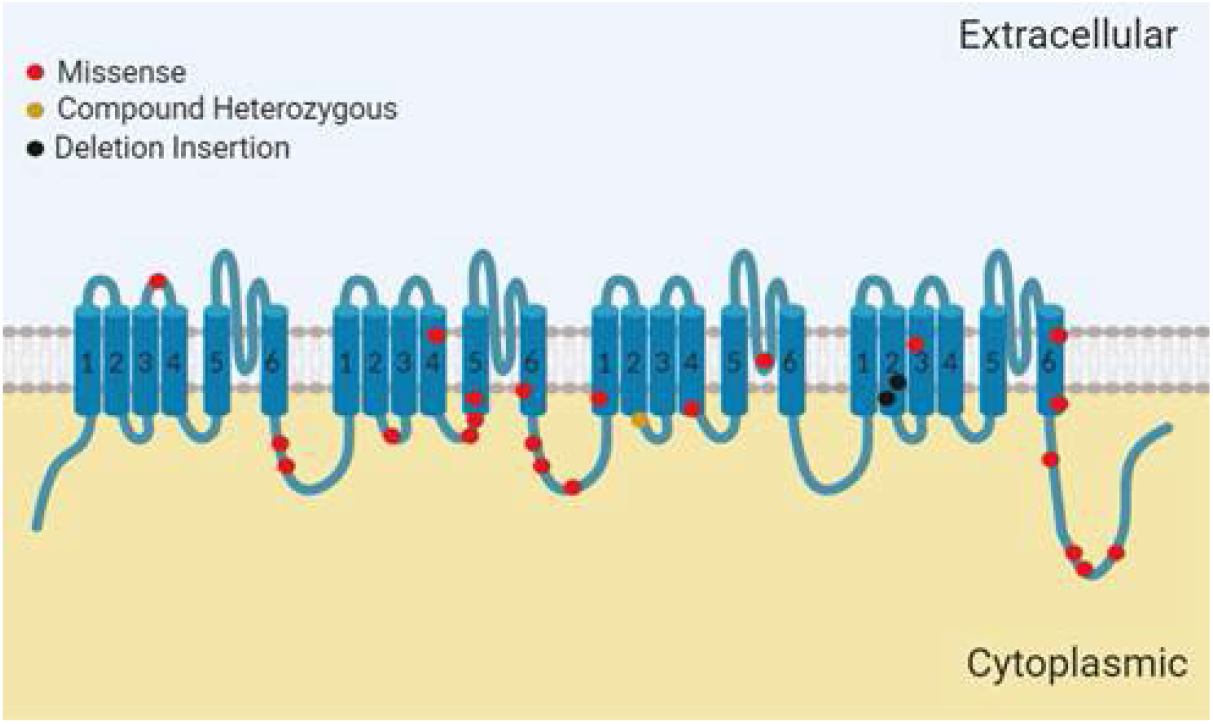
Two-dimensional structure of *SCN2A* α subunit with variants identified in this cohort.

**Supplemental Figure 2:**
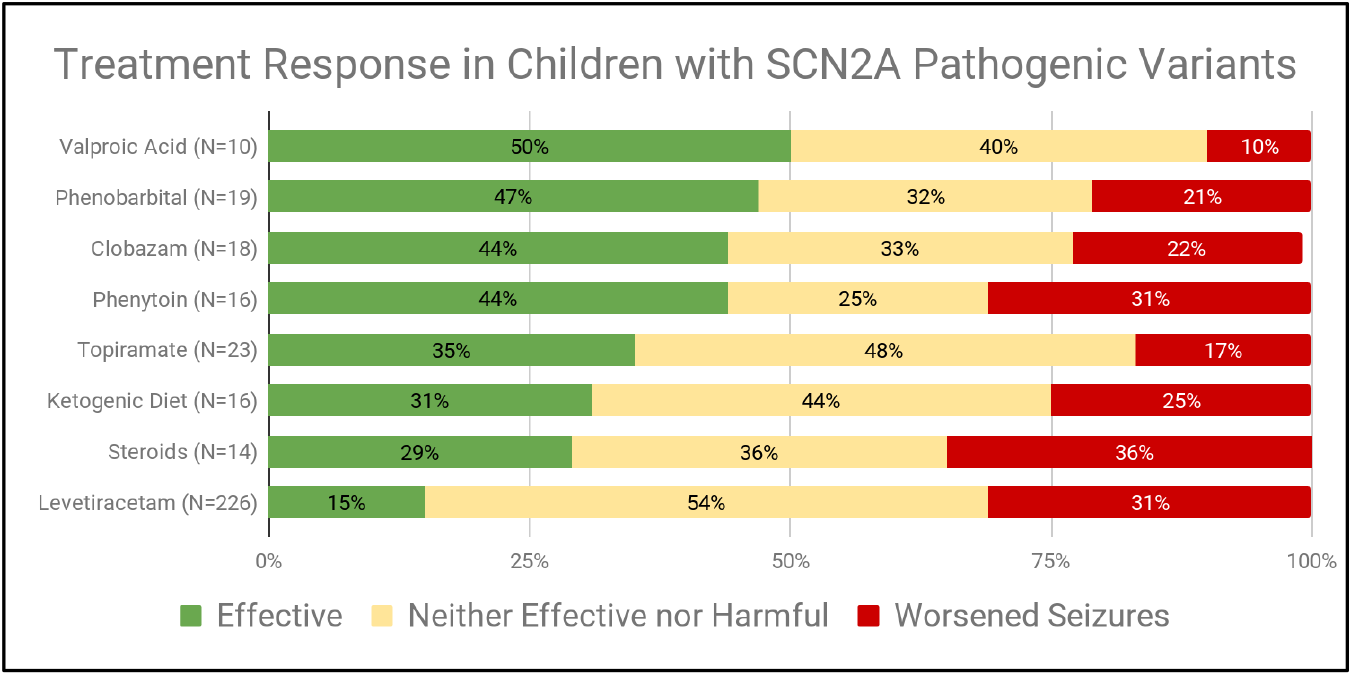
Parent-reported responses to previously trialed anti-epileptic treatments in *SCN2A* participants.

**Supplemental Table 1:** Summary of the Reported Single Most Effective Treatment for their Child’s Seizures (N=19; excluding other medications)

| **Treatment** | **Participants (No.)** |
| --- | --- |
| Phenytoin | 4 |
| Topiramate | 3 |
| Clobazam | 2 |
| Ketogenic Diet | 2 |
| Valproic Acid | 1 |
| Levetiracetam | 1 |
| Carbamazepine | 1 |
| Lamotrigine | 1 |
| Other | 4 |

**Supplemental Table 2:** Count of Treatments Reported to be Effective in Combination with Another Medication (excluding unreported treatments)

| **Treatment** | **Participants (No.)** |
| --- | --- |
| Topiramate | 6 |
| Clobazam | 5 |
| Lamotrigine | 4 |
| Phenobarbital | 4 |
| Carbamazepine | 3 |
| Valproic Acid | 3 |
| Ketogenic Diet | 3 |
| Lacosamide | 2 |
| Levetiracetam | 2 |
| Phenytoin | 2 |
| Ethosuximide | 1 |
| Gabapentin | 1 |
| Perampanel | 1 |
| Rufinamide | 1 |
| Steroids | 1 |
| Vigabatrin | 1 |
| Vagal Nerve Stimulator | 1 |
| Other | 7 |

## Notes

### Competing Interest Statement

All authors have completed the ICMJE uniform disclosure form at www.icmje.org/coi_disclosure.pdf and declare: no support from any organisation for the submitted work.

### Funding Statement

This study did not receive any funding

### Author Declarations

Approved by the Institutional Review Board (IRB) of Ann and Robert H. Lurie Childrens Hospital of Chicago with low-risk designation.

